# Evaluation of a DNA methylation-based measure of chronic inflammation in two generations of adults in metropolitan Cebu, Philippines

**DOI:** 10.1101/2025.03.25.25324477

**Authors:** Thomas W. McDade, Sofia Carrera, Calen P. Ryan, David Burgner, Linda S. Adair, Nanette R. Lee, Delia B. Carba, Julia L. MacIsaac, Kristy Dever, Michael S. Kobor, Christopher W. Kuzawa

## Abstract

**Objectives:** Proxy measures of chronic inflammation derived from DNA methylation (DNAm) data have emerged as promising predictors of cardiometabolic disease risk in high income countries. This study investigates the performance of a recently validated DNAm-based measure of C-reactive protein (DNAm-CRP) in two generations of adults in the Philippines to evaluate its utility in lower and middle income settings experiencing high levels of endemic infections as well as rising rates of chronic degenerative diseases.

**Methods:** DNAm-CRP was calculated from 1,468 CpG sites on the Infinium MethylationEPIC v1.0 array applied to genomic DNA from leukocytes in young adults (N=1,665; 20-22 years) and older women (N=1,070; 35-68 years). C-reactive protein was determined in plasma using a high sensitivity immunoturbidimetric assay. Pearson correlation and least squares regression were implemented to evaluate the strength of association between DNAm-CRP and plasma CRP, and to investigate patterns of association between DNAm-CRP and established predictors of chronic inflammation.

**Results:** For younger adults, the correlation between DNAm-CRP and log-transformed CRP was 0.41, and DNAm-CRP explained 17.2% of the variance in CRP. For older women, the correlation was 0.47, with 22.7% explained variance in CRP. For both cohorts larger waist circumference was associated with higher DNAm-CRP. The presence of infectious symptoms at the time of blood collection and leukocyte composition were both significant predictors of DNAm-CRP.

**Conclusions:** In two generations of adults in the Philippines, we document strong correlations between DNAm-CRP and plasma CRP. DNAm-CRP may be a useful tool for research on chronic inflammation across a range of epidemiological and ecological settings globally, but future applications should consider how recent infections and the distribution of leukocyte subsets may confound or mediate associations of interest.

## INTRODUCTION

Chronic low-grade inflammation is an independent risk factor for incident heart disease, stroke, type 2 diabetes, and other chronic degenerative diseases of aging, and a strong predictor of all-cause mortality [1, 2]. C-reactive protein (CRP) is an acute phase protein and established blood-based biomarker of systemic inflammation that is widely used clinically and in large-scale epidemiological and population-based studies [3–5]. However, CRP concentrations increase rapidly in response to infection or injury [6], potentially obscuring the “signal” of chronic inflammation when CRP is measured at a single timepoint. Acute elevations of CRP pose particular challenges to measurement in epidemiological contexts where infections are common [7, 8].

The convergence of genome-wide DNA methylation (DNAm) arrays and novel bioinformatic approaches has generated new opportunities for identifying measures of biological aging and circulating proteins in the epigenome [9, 10]. The methylation of DNA involves reversible binding of methyl groups to cytosine residues, primarily in the context of CpG dinucleotides. Environmental exposures and other genomic processes influence the pattern of DNAm, which in turn can have effects on the regulation of gene expression [11]. DNAm is therefore a promising candidate for understanding the mechanisms through which contexts and behaviors can shape health trajectories over the life course. This is particularly the case for inflammation, since routine blood sampling provides access to important tissues (i.e., leukocytes) that regulate inflammation. Genome-wide DNAm data have been used to develop several surrogate measures of inflammation that correlate with circulating CRP concentrations and predict a range of cardiometabolic and aging-related health outcomes [12–16]. Associations are generally as strong, or stronger, than associations with CRP, and DNAm-based measures of chronic inflammation appear to be relatively stable over time and less sensitive than CRP to acute fluctuation [12, 17].

A limitation of current research on DNAm-based inflammation scores is that it is based almost exclusively on studies of older adults in Canada, Europe, and the United States. The extent to which DNAm-based measures of inflammation developed in epidemiologic contexts defined by low levels of infectious disease and high rates of overweight and obesity will translate to other settings globally is not known. Addressing this question is important since approximately 75% of the global burden of death and disability from cardiovascular diseases occurs in low-and middle-income countries [18, 19], many of which are enduring high levels of endemic infections as well as rising rates of chronic degenerative diseases [20, 21].

The objective of this study is to investigate the performance of a recently validated DNAm-based surrogate measure of CRP (DNAm-CRP) in two generations of adults in the Philippines. Despite recent gains in educational opportunities, household income, and life expectancy, the Philippines remains a lower-middle income nation with rising burdens of cardiometabolic disease against a background of high rates of infectious disease [22–24]. Communicable, maternal, perinatal, and nutritional conditions account for approximately one in four deaths [25].

We selected DNAm-CRP because it explains more variance in CRP than prior measures, and because test cohorts included individuals of diverse ancestry, as well as age groups from infancy through the elderly [17]. The measure represents the optimal weighted linear combination of CpG sites that predict circulating CRP concentrations, and it has been tested in six cohort studies comprising approximately 25,000 individuals. It has high within-individual stability across time, and is positively associated with risk for cardiovascular disease, diabetes, and all-cause mortality. In addition, the DNAm-CRP is constructed from CpG sites that are measured in both the Illumina 450k and EPIC v1.0 arrays, therefore providing backward compatibility with earlier studies. We evaluate strength of correlation with CRP in two generations of Filipinos: Young adults and their mothers, both of whom are participants in an ongoing multi-generation cohort study. We document strong correlations between DNAm-CRP and circulating CRP in both cohorts, as well as associations with established predictors of chronic inflammation. We also find that the distribution of leukocyte subsets is a strong predictor of DNAm-CRP, highlighting the importance of considering cell composition in analyses of associations among environmental exposures, DNAm-based inflammation scores, and health outcomes.

## MATERIALS AND METHODS

### Participants and data collection

The Cebu Longitudinal Health and Nutrition Survey (CLHNS) is a multi-generation prospective cohort study initiated in 1983 with the recruitment of 3,327 pregnant women representative of the childbearing population in Metropolitan Cebu, Philippines [23]. Data have been collected from the women and their children (N=3,080 singleton live births) across multiple survey waves, including an in-home survey in 2005 that collected overnight fasting venous blood samples for biomarker and genetic analyses. All the children were born in 1983-84, and were 20-22 years old during the 2005 survey. Mothers ranged in age from 35 to 68 years (we refer to this cohort as “older women” from this point forward, to distinguish them from the cohort of younger adults).

Blood was drawn into EDTA-coated vacutainer tubes, transported in coolers with ice packs, and centrifuged to separate plasma and leukocytes prior to freezing at-70°C. Samples were express shipped on dry ice and stored at-80°C prior to analysis. Concentrations of CRP were determined in plasma using a high sensitivity immunoturbidimetric method (Synchron LX20, lower detection limit: 0.1 mg/L). Interviewers implemented standard procedures to measure waist circumference above the umbilicus over light clothing [26]. All data were collected under conditions of informed consent with institutional review board approval from the University of North Carolina at Chapel Hill and Northwestern University.

The 2005 survey included 1,885 young adults, 1,731 of whom provided complete data for these analyses. Women who were pregnant (n=66) at the time of the survey were not included due to the effect of pregnancy on inflammation, resulting in a final sample size of 1,665.

N=2,018 older women were interviewed in 2005, with a final sample size of 1,070. All available DNA samples were analyzed for the young adults, whereas a random subset of approximately half of the available DNA samples collected in 2005 were analyzed for older women. Four older women who were pregnant at the time of blood collection were not included.

*DNA methylation and calculation of DNAm-CRP.* Genomic DNA from white blood cells was treated with sodium bisulfite (EZ DNA Methylation kit, Zymo Research), and converted DNA was applied to the Infinium MethylationEPIC v1.0 array following manufacturer’s standard protocol (Illumina Inc., San Diego, CA, USA). Samples were randomized across sampling units, and technical replicates were included to confirm consistent results across arrays [27]. Data were noob (normal-exponential out-of-band) normalized using the R package *minfi* before further analysis [28–30].

DNAm-CRP was generated using a weighted linear combination of 1,468 CpG sites previously identified as the optimal predictors of log-transformed CRP in elastic net regression [17] (original code available at https://doi.org/10.5281/zenodo.10154736). Prior to statistical analyses, DNAm-CRP was standardized (mean = 0, SD = 1) to facilitate interpretation. The distribution of white blood cell subsets was determined from the noob-corrected data using the FlowSorted.BloodExtended.EPIC (version 0.99.0) R package, a cell deconvolution algorithm developed for EPICv1 DNA methylation arrays [31]. Due to collinearity in cell proportion data (i.e., total cell proportions sum to 1.0), we generated four principal component scores that captured 92.8% of the variance in cell composition for young adults, and 91.3% for older women.

### Data analysis

Pearson correlation and least squares regression were implemented to evaluate the strength of association between DNAm-CRP and log10-transformed CRP. Prior to log transformation, CRP values below the assay lower detection limit were assigned a value of 0.01 mg/L to retain all observations, following prior validation studies [17]. Proportion of CRP variance explained by DNAm-CRP was evaluated with incremental R^2^ estimates that subtracted the adjusted R^2^ value attributed to age and sex from the adjusted R^2^ in a regression model that included age, sex, and DNAm-CRP. DNAm-CRP was also analyzed as the dependent variable in regression models with established predictors of chronic inflammation. Analyses were conducted with Stata for Windows, version 15 (StataCorp, College Station, TX), with p<0.05 set as the criterion for statistical significance.

Predictors included sex (assigned at birth), age, smoking (indicator variable for daily smoker), waist circumference, years of formal education, household assets, household microbial exposure, and infection at the time of blood collection. Household assets provide a more stable measure of economic status than income in this setting [32], and a summary variable was constructed from the following items: home ownership, electricity in the home, type of housing material, and ownership of items such as air conditioner, television, refrigerator, or car.

Following prior analyses with this dataset, a variable was constructed to estimate the level of exposure to infectious microbes in the household [33]. Separate variables were constructed for both cohorts based on a sum of the following: household density (number of persons/number of rooms), interviewer rating of presence of garbage and fecal contamination in the home and in the neighborhood, and source of drinking water (closed sources: bottled, piped municipal supply, closed well with pump, vs. open sources: uncovered well, spring, river, rain). Current infection (0, 1) was defined by participant report of any of the following symptoms at the time of blood collection: runny nose, cough, fever, diarrhea, sore throat, as well as the more general categories of “flu,” “cold,” and “sinusitis”.

The analysis plan was designed to be comparable to the validation analyses implemented by Hillary et al. [17]. However, we implemented a series of follow up analyses to determine if results were sensitive to acute inflammation at the time of blood collection, and to censoring of the CRP distribution (values <0.1 mg/L), both of which occurred more frequently in our datasets. Spearman’s rank correlations and tobit regressions were both applied to logCRP data to evaluate robustness of results to deviations from assumptions of normality in the logCRP distribution.

We also replicated analyses excluding participants with infectious symptoms at the time of blood collection, and with CRP>10 mg/L, a commonly used cut-point to identify acute inflammation [4].

## RESULTS

Table 1 presents descriptive statistics for both cohorts. The bivariate correlation between DNAm-CRP and log-transformed CRP was 0.47 and 0.41 for older females and young adults, respectively. When considered separately, the Pearson correlation among young adults was similar for females (0.42) and males (0.41). Scatterplots demonstrate the positive associations between DNAm-CRP and plasma CRP, but also document a wide range of DNAm-CRP values for participants with very low CRP (Figure 1). Since a substantial proportion of both samples have concentrations of CRP<0.1 mg/L (13.2% of older females, 33.9% of young adults), we calculated correlations excluding observations with undetectable CRP. Results were comparable for older females (R=0.47) and young adults (R=0.42). For older females, DNAm-CRP explained 22.7% of the incremental variance in CRP after adjusting for age. For younger adults, DNAm-CRP explained 17.2% of the incremental variance in CRP adjusting for age and sex.

**Figure 1.**
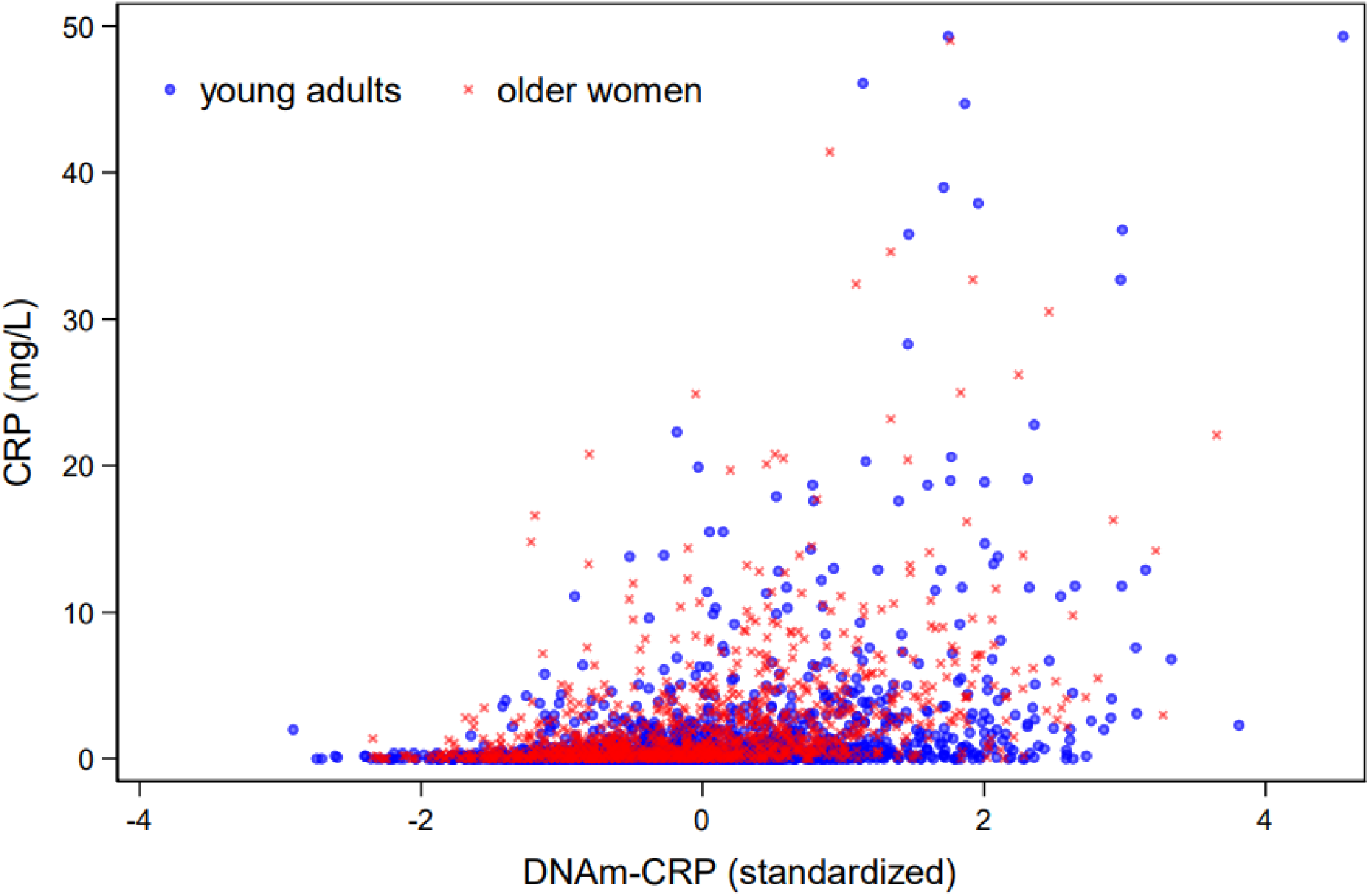
Bivariate association between DNAm-CRP and plasma CRP for younger adults and older women in the Philippines. For scaling purposes, nine observations with plasma CRP>50 mg/L are not included in the figure (CRP and DNAm-CRP values, respectively, for these observations are as follows: 55.4, 2.3; 55.7, 3.9; 58.5, 2.3; 64.9, 2.2; 67.9, 0.9; 74.0, 1.8; 79.0, 2.5; 85.9, 2.84; 122.6, 5.3).

**Table 1.**
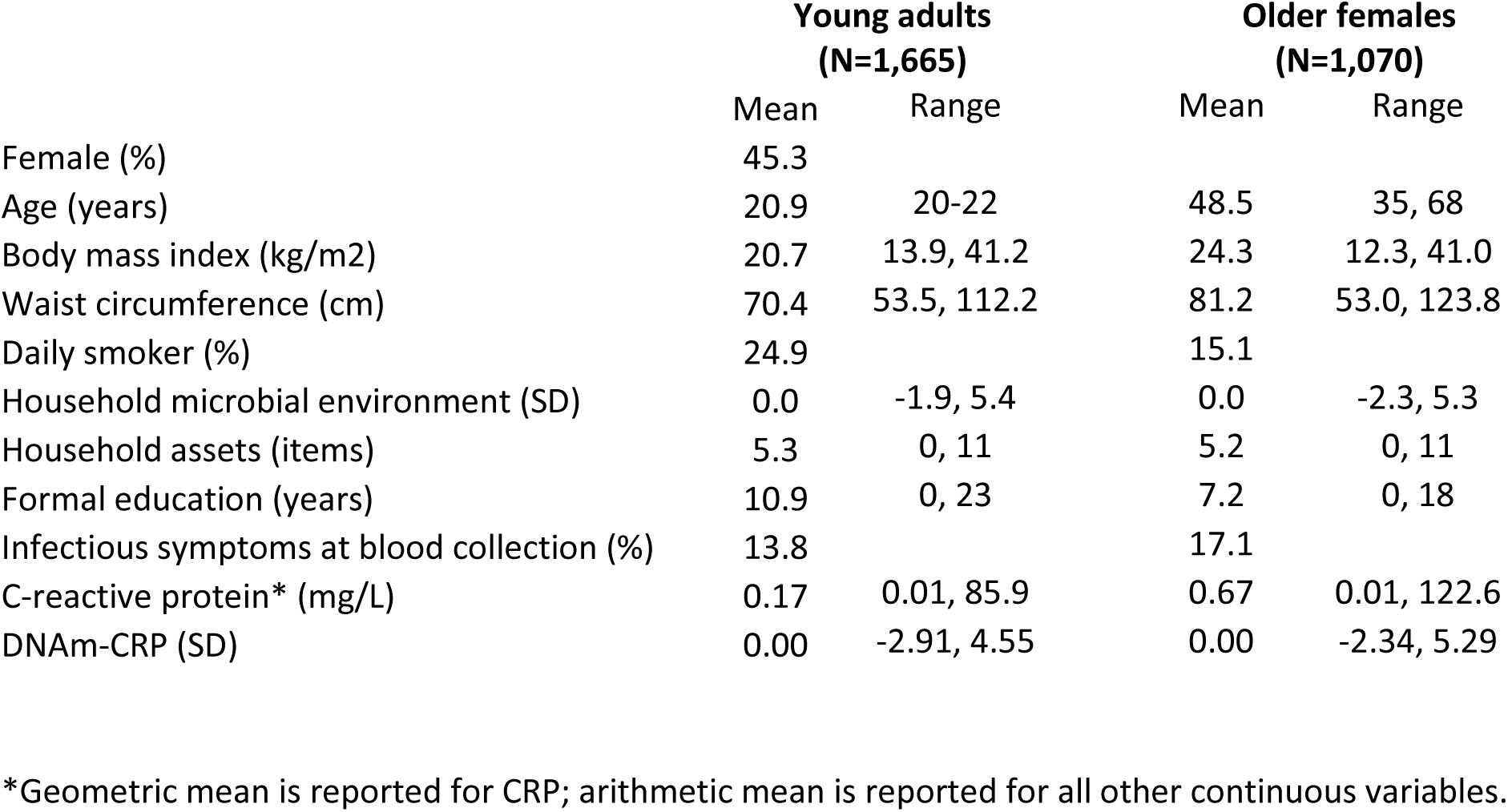
Descriptive statistics for the two study samples, measured in 2005.

|  | **Young adults**<br>**(N=1,665)** |  | **Older females**<br>**(N=1,070)** |  |
| --- | --- | --- | --- | --- |
|  | Mean | Range | Mean | Range |
| Female (%) | 45.3 |  |  |  |
| Age (years) | 20.9 | 20-22 | 48.5 | 35, 68 |
| Body mass index (kg/m<sup>2</sup>) | 20.7 | 13.9, 41.2 | 24.3 | 12.3, 41.0 |
| Waist circumference (cm) | 70.4 | 53.5, 112.2 | 81.2 | 53.0, 123.8 |
| Daily smoker (%) | 24.9 |  | 15.1 |  |
| Household microbial environment (SD) | 0.0 | -1.9, 5.4 | 0.0 | -2.3, 5.3 |
| Household assets (items) | 5.3 | 0, 11 | 5.2 | 0, 11 |
| Formal education (years) | 10.9 | 0, 23 | 7.2 | 0, 18 |
| Infectious symptoms at blood collection (%) | 13.8 |  | 17.1 |  |
| C-reactive protein* (mg/L) | 0.17 | 0.01, 85.9 | 0.67 | 0.01, 122.6 |
| DNAm-CRP (SD) | 0.00 | -2.91, 4.55 | 0.00 | -2.34, 5.29 |

\*Geometric mean is reported for CRP; arithmetic mean is reported for all other continuous variables.

In bivariate (Table S1) as well as multivariate analyses (Table 2), many established predictors of chronic inflammation were associated with DNAm-CRP. For both cohorts larger waist circumference was significantly associated with higher DNAm-CRP. Young adult females had significantly higher scores than males. Level of education was negatively associated with DNAm-CRP for younger adults, but not for older females. Age did not predict DNAm-CRP for younger adults, which is to be expected given the narrow age range of this one-year birth cohort. For older females, there was a non-linear association with age such that DNAm-CRP trended downward, but was higher for women approximately 60 years and older. Presence of infectious symptoms at the time of blood collection was associated with higher DNAm-CRP for both cohorts.

**Table 2.** Predictors of DNAm-CRP in least squares regression models for younger adults and older females, with and without adjustment for cell composition.

|  | Young adults |  |  |  |  |  | Older females |  |  |  |  |  |
| --- | --- | --- | --- | --- | --- | --- | --- | --- | --- | --- | --- | --- |
|  | B | SE | P | B | SE | P | B | SE | P | B | SE | P |
| Age (years) | 0.011 | 0.068 |  | -0.028 | 0.060 |  | -0.171 | 0.065 | ** | -0.111 | 0.062 | + |
| Age<sup>2</sup> |  |  |  |  |  |  | 0.002 | 0.001 | * | 0.001 | 0.001 | + |
| Sex (female = 1) | 0.461 | 0.054 | *** | 0.549 | 0.052 | *** |  |  |  |  |  |  |
| Current smoker (0, 1) | -0.093 | 0.062 |  | -0.006 | 0.056 |  | 0.093 | 0.084 |  | 0.048 | 0.081 |  |
| Household microbial exposure (SD) | 0.041 | 0.024 | + | 0.058 | 0.021 | ** | -0.001 | 0.033 |  | 0.016 | 0.031 |  |
| Household assets (items) | -0.027 | 0.013 | + | -0.012 | 0.012 |  | -0.007 | 0.019 |  | -0.001 | 0.018 |  |
| Education (years) | -0.023 | 0.007 | ** | -0.019 | 0.006 | ** | 0.000 | 0.009 |  | 0.002 | 0.008 |  |
| Waist circumference (cm) | 0.029 | 0.003 | *** | 0.027 | 0.003 | *** | 0.026 | 0.003 | *** | 0.027 | 0.003 | *** |
| Infectious symptoms (0, 1) | 0.356 | 0.067 | *** | 0.296 | 0.060 | *** | 0.242 | 0.078 | ** | 0.231 | 0.074 | ** |
| PC1 (SD) |  |  |  | -0.367 | 0.021 | *** |  |  |  | -0.296 | 0.028 | *** |
| PC2 (SD) |  |  |  | -0.229 | 0.021 | *** |  |  |  | -0.133 | 0.029 | *** |
| PC3 (SD) |  |  |  | 0.052 | 0.021 | ** |  |  |  | 0.074 | 0.028 | ** |
| PC4 (SD) |  |  |  | 0.159 | 0.022 | *** |  |  |  | -0.026 | 0.028 |  |
| Constant | -2.121 | 1.459 |  | -1.298 | 1.289 |  | 2.312 | 1.628 |  | 0.474 | 1.550 |  |
| Model R<sup>2</sup> (adjusted) | 0.101 |  |  | 0.301 |  |  | 0.088 |  |  | 0.191 |  |  |

Note: +p<0.10; \*p<0.05; \*\*p<0.01; \*\*\*p<0.001;
Note: PC1, PC2, PC3 and PC4 are principal component scores capturing variance in white blood cell composition.

Leukocyte composition was a strong predictor of DNAm-CRP. In bivariate analyses, proportions of CD4 T lymphocytes, CD8 T lymphocytes, B lymphocytes, and NK cells were negatively correlated with DNAm-CRP in young adults and older females. Monocyte and granulocyte proportions were positively associated with DNAm-CRP for both cohorts (Table S1). When variables representing leukocyte composition were added to regression models, the proportion of explained variance in DNAm-CRP increased from 8.9 to 19.1% for older females, and from 10.0 to 30.1% for younger adults. Coefficients for other predictors were largely unchanged, with the exception of household microbial exposure which was positively, and significantly, associated with DNAm-CRP for younger adults after adjusting for cell composition. Associations with age were attenuated for older females following leukocyte adjustment.

In sensitivity analyses, Spearman’s rank correlations between DNAm-CRP and logCRP were similar to Pearson correlations for older women (rho=0.49), and were not substantially altered when eliminating participants with current infection (rho=0.49) or CRP>10 mg/L (rho=0.46). The pattern was similar for younger adults (rho=0.39), with slightly reduced correlations when excluding observations with current infection (rho=0.36) or CRP>10 mg/L (rho=0.34). As with ordinary least squares results, tobit regression models indicated that DNAm-CRP was a strong and significant predictor of logCRP for older women (B=0.467, SE=0.028, p<0.001) and younger adults (B=0.578, SE=0.035, p<0.001). However, estimates of explained variance in CRP were lower: Based on the incremental pseudo R^2^, DNAm-CRP explained 8.5% of the variance in CRP for older women, and 5.4% for younger adults.

## DISCUSSION

Chronic systemic inflammation is an established risk factor for degenerative diseases of aging, but acute fluctuations in inflammatory biomarkers can make it difficult to accurately quantify chronic inflammation from a single blood draw. DNAm-based predictors of CRP have emerged as relatively stable markers of chronic inflammation for adults living in high income countries with low rates of infectious disease, but their performance in a broader global context has not been evaluated. In two generations of adults in the Philippines, a lower-middle income country experiencing dual burdens of infectious and chronic degenerative diseases, we document strong correlations between DNAm-CRP and circulating concentrations of CRP, as well as significant associations with established determinants of chronic inflammation, both of which suggest that DNAm-CRP may be a useful tool for research into the causes and consequences of chronic inflammation across a range of epidemiological and ecological settings globally.

We focused our analyses on a previously validated DNAm-based predictor of circulating CRP, with correlations that range from 0.27 to 0.53 in cohorts of younger and older adults [17]. The Avon Longitudinal Study of Parents and Children (ALSPAC) provides a useful comparison for our study in that it includes DNAm-CRP and CRP measures for young adults (mean age 24.5 years) and their mothers (mean age 47.8 years), with correlations of 0.37 and 0.39, respectively [17]. The Health for Life in Singapore study is geographically more proximate to our study site in the Philippines and includes participants of Chinese, Malay, and Indian ancestry (mean ages 51.3-55.4 years), with correlations of 0.39, 0.45, and 0.53, respectively [17]. Overall, we find that the strength of correlation between DNAm-CRP and plasma CRP in younger and older adults in the Philippines is within the range of what has been reported in other validation cohorts.

The validity of DNAm-CRP as a measure of chronic inflammation in this context is further supported by positive associations with waist circumference in both generations of participants. Central body fat contributes to chronic inflammation, even when average levels of adiposity are relatively low, as is the case in both our young adult and older female participants [34, 35]. It is also notable that associations between DNAm-CRP and waist circumference, and between DNAm-CRP and plasma CRP, are evident even though the overall level of chronic inflammation is quite low with mean CRP of 0.17 and 0.67 mg/L for young adults and older females in our study, respectively. In comparison, mean CRP is 1.7 mg/L for young adults in ALSPAC, and 1.8 mg/L for their mothers [17].

It is also interesting to note that the presence of infectious symptoms at the time of blood collection was associated with significantly higher DNAm-CRP scores for both cohorts in our study. Elevations in circulating CRP are to be expected with current or recent infection as part of the acute phase response [6]. However, a proposed advantage of DNAm-CRP and other methylation-based proxies of inflammation is that they provide more accurate measures of chronic activity because they are less sensitive to acute fluctuation [17]. Our results suggest additional research is needed to determine the temporal stability of DNAm-CRP in settings where infectious exposures are common, particularly since *in vitro* experiments have documented differential methylation of CpG sites in monocytes following exposure to low physiological doses of CRP [12].

Leukocyte proportions were strong predictors of DNAm-CRP, and explained substantially more variance in DNAm-CRP than waist circumference and other common predictors of chronic inflammation. The distribution of leukocyte subsets varies considerably across individuals, and prior research has demonstrated that exposures during development can influence cell composition in adulthood [27, 36, 37]. Similarly, peripheral blood leukocyte composition changes with age, and is associated with incident cardiovascular disease and mortality risk [38–40]. It is therefore essential that future applications of DNAm-CRP and other DNAm-based measures of inflammation carefully consider the extent to which leukocyte fractions may mediate, or confound, associations of interest.

Our findings suggest that DNAm-based measures may be useful tools for research on inflammation and aging globally. Based on studies in affluent countries with high levels of overweight and obesity, the concept of “inflammaging” points toward chronic, dysregulated inflammation as both a cause and consequence of degenerative processes that lead to age-related increases in morbidity and mortality [41]. However, it is not clear if “inflammaging” generalizes to lower-and middle-income settings where endemic infectious diseases mobilize acute inflammatory responses but may also establish immunoregulatory networks early in development that reduce burdens of chronic inflammation in adulthood [33, 42, 43]. Measures of chronic inflammation that are not obscured by acute phase responses can advance our understanding of how inflammation may contribute to chronic degenerative diseases globally, and guide efforts at prevention that are most effective in a particular ecological and epidemiological setting.

Limitations of our study include single timepoint measures of DNAm-CRP and CRP, which prevent us from comparing temporal stability of both measures. In addition, the older cohort includes only females. Even though we find no sex difference in patterns of association in the young adult cohort, additional research should be conducted with older males. It is also important to note that DNAm-CRP correlates with circulating CRP, but patterns of causality are not known. Additional research is needed to determine whether acute or chronic elevations in CRP may modify the methylation status of CpG sites in the measure, and/or whether modifications to these sites influence the production of CRP.

## Data Availability

The data that support the findings of this study are available to qualified users following approval of a data use agreement. The data are not publicly available due to privacy or ethical restrictions.

## ACKNOWLEDGEMENTS

We thank the researchers at the USC-Office of Population Studies Foundation, Inc., University of San Carlos, Cebu City, Philippines, for their role in the study design and data collection, and the study participants, who generously provided their time. Research reported in this publication was supported by the National Institute on Aging of the National Institutes of Health (R01 AG061006R21). The content is solely the responsibility of the authors and does not necessarily represent the official views of the National Institutes of Health. TM gratefully acknowledges financial support from CIFAR as a fellow of the Child and Brain Development program. This research was enabled, in part, by the FlowSorted.BloodExtended.EPIC software packages developed at Dartmouth College, which are subject to the licensing terms made available by Dartmouth Technology Transfer.

## REFERENCES

1. Furman, D., J. Campisi, E. Verdin, P. Carrera-Bastos, S. Targ, C. Franceschi, L. Ferrucci, D.W. Gilroy, A. Fasano, and G.W. Miller, Chronic inflammation in the etiology of disease across the life span. Nature Medicine, 2019. 25(12): p. 1822–1832.

2. Proctor, M.J., D.C. McMillan, P.G. Horgan, C.D. Fletcher, D. Talwar, and D.S. Morrison, Systemic inflammation predicts all-cause mortality: a Glasgow inflammation outcome study. PLoS One, 2015. 10(3): p. e0116206.

3. Emerging Risk Factors Collaboration, C-reactive protein concentration and risk of coronary heart disease, stroke, and mortality: an individual participant meta-analysis. The Lancet, 2010. 375(9709): p. 132-140.

4. Pearson, T.A., G.A. Mensah, R.W. Alexander, J.L. Anderson, R.O. Cannon, M. Criqui, Y.Y. Fadl, S.P. Fortmann, Y. Hong, G.L. Myers, N. Rifai, S.C. Smith, K. Taubert, R.P. Tracy, and F. Vinicor, Markers of inflammation and cardiovascular disease: Application to clinical and public health practice. Circulation, 2003. 107: p. 499–511.

5. McDade, T.W., J.M. Meyer, S.M. Koning, and K.M. Harris, Body mass and the epidemic of chronic inflammation in early mid-adulthood. Social Science and Medicine, 2021. 281: p. 114059.

6. Ballou, S.P. and I. Kushner, C-reactive protein and the acute phase response. Advances in Internal Medicine, 1992. 37: p. 313–336.

7. Bogaty, P., G.R. Dagenais, L. Joseph, L. Boyer, A. Leblanc, P. Belisle, and J.M. Brophy, Time variability of C-reactive protein: implications for clinical risk stratification. PLoS One, 2013. 8(4): p. e60759.

8. McDade, T.W., P.S. Tallman, F.C. Madimenos, M.A. Liebert, T.J. Cepon, L.S. Sugiyama, and J.J. Snodgrass, Analysis of variability of high sensitivity C-reactive protein in lowland Ecuador reveals no evidence of chronic low-grade inflammation. American Journal of Human Biology, 2012. 24: p. 675–81.

9. Gadd, D.A., R.F. Hillary, D.L. McCartney, S.B. Zaghlool, A.J. Stevenson, Y. Cheng, C. Fawns-Ritchie, C. Nangle, A. Campbell, and R. Flaig, Epigenetic scores for the circulating proteome as tools for disease prediction. eLife, 2022. 11: p. e71802.

10. Ryan, C.P., “Epigenetic clocks”: Theory and applications in human biology. American Journal of Human Biology, 2021. 33(3): p. e23488.

11. Aristizabal, M.J., I. Anreiter, T. Halldorsdottir, C.L. Odgers, T.W. McDade, A. Goldenberg, S. Mostafavi, M.S. Kobor, E.B. Binder, and M.B. Sokolowski, Biological embedding of experience: a primer on epigenetics. Proceedings of the National Academy of Sciences, 2020. 117(38): p. 23261–23269.

12. Verschoor, C.P., C. Vlasschaert, M.J. Rauh, and G. Paré, A DNA methylation based measure outperforms circulating CRP as a marker of chronic inflammation and partly reflects the monocytic response to long-term inflammatory exposure: A Canadian Longitudinal Study on Aging analysis. Aging Cell, 2023. 22(7): p. e13863.

13. Conole, E.L., A.J. Stevenson, S. Muñoz Maniega, S.E. Harris, C. Green, M.d.C. Valdés Hernández, M.A. Harris, M.E. Bastin, J.M. Wardlaw, and I.J. Deary, DNA methylation and protein markers of chronic inflammation and their associations with brain and cognitive aging. Neurology, 2021. 97(23): p. e2340–e2352.

14. Ligthart, S., C. Marzi, S. Aslibekyan, M.M. Mendelson, K.N. Conneely, T. Tanaka, E. Colicino, L.L. Waite, R. Joehanes, and W. Guan, DNA methylation signatures of chronic low-grade inflammation are associated with complex diseases. Genome Biology, 2016. 17: p. 1–15.

15. Wielscher, M., P.R. Mandaviya, B. Kuehnel, R. Joehanes, R. Mustafa, O. Robinson, Y. Zhang, B. Bodinier, E. Walton, and P.P. Mishra, DNA methylation signature of chronic low-grade inflammation and its role in cardio-respiratory diseases. Nature Communications, 2022. 13(1): p. 2408.

16. Meier, H.C., C. Mitchell, T. Karadimas, and J.D. Faul, Systemic inflammation and biological aging in the Health and Retirement Study. GeroScience, 2023. 45(6): p. 3257–3265.

17. Hillary, R.F., H.K. Ng, D.L. McCartney, H.R. Elliott, R.M. Walker, A. Campbell, F. Huang, K. Direk, P. Welsh, and N. Sattar, Blood-based epigenome-wide analyses of chronic low-grade inflammation across diverse population cohorts. Cell Genomics, 2024. 4(5).

18. Gurven, M.D. and D.E. Lieberman, WEIRD bodies: mismatch, medicine and missing diversity. Evolution and Human Behavior, 2020. 41(5): p. 330–340.

19. Di Cesare, M., P. Perel, S. Taylor, C. Kabudula, H. Bixby, T.A. Gaziano, D.V. McGhie, J. Mwangi, B. Pervan, and J. Narula, The heart of the world. Global Heart, 2024. 19(1): p. 11.

20. United Nations, Population Division, World Population Prospects: The 2015 Revision, Volume I: Comprehensive Tables (ST/ESA/SER.A/379). 2015.

21. Boutayeb, A., The double burden of communicable and non-communicable diseases in developing countries. Transactions of the Royal Society of Tropical Medicine and Hygiene, 2006. 100(3): p. 191–199.

22. World Health Organization, The World Health Report 2004 - Changing History. 2004, World Health Organization.

23. Adair, L.S., B.M. Popkin, J.S. Akin, D.K. Guilkey, S. Gultiano, J. Borja, L. Perez, C.W. Kuzawa, T. McDade, and M.J. Hindin, Cohort profile: the Cebu Longitudinal Health and Nutrition Survey. Int J Epidemiol, 2011. 40(3): p. 619–25.

24. Badana, A.N. and R. Andel, Aging in the Philippines. The Gerontologist, 2018. 58(2): p. 212–218.

25. World Health Organization. Health data overview for the Republic of the Philippines. 2021 [cited 2025 March 17]; Available from: https://data.who.int/countries/608.

26. Lohman, T.G., A.F. Roche, and R. Martorell, Anthropometric Standardization Reference Manual. 1988, Champaign, IL: Human Kinetics Books.

27. McDade, T.W., C.P. Ryan, L.S. Adair, N.R. Lee, D.B. Carba, J.L. MacIsaac, K. Dever, P. Atashzay, M.S. Kobor, and C.W. Kuzawa, Association between infectious exposures in infancy and epigenetic age acceleration in young adulthood in metropolitan Cebu, Philippines. American Journal of Human Biology, 2023. 35(11): p. e23948.

28. Triche, T.J., Jr, D.J. Weisenberger, D. Van Den Berg, P.W. Laird, and K.D. Siegmund, Low-level processing of Illumina Infinium DNA Methylation BeadArrays. Nucleic Acids Research, 2013. 41(7): p. e90–e90.

29. Aryee, M.J., A.E. Jaffe, H. Corrada-Bravo, C. Ladd-Acosta, A.P. Feinberg, K.D. Hansen, and R.A. Irizarry, Minfi: a flexible and comprehensive Bioconductor package for the analysis of Infinium DNA methylation microarrays. Bioinformatics, 2014. 30(10): p. 1363–1369.

30. Fortin, J.-P., T.J. Triche, Jr, and K.D. Hansen, Preprocessing, normalization and integration of the Illumina HumanMethylationEPIC array with minfi. Bioinformatics, 2016. 33(4): p. 558–560.

31. Salas, L.A., Z. Zhang, D.C. Koestler, R.A. Butler, H.M. Hansen, A.M. Molinaro, J.K. Wiencke, K.T. Kelsey, and B.C. Christensen, Enhanced cell deconvolution of peripheral blood using DNA methylation for high-resolution immune profiling. Nature Communications, 2022. 13(1): p. 761.

32. Howe, L.D., B. Galobardes, A. Matijasevich, D. Gordon, D. Johnston, O. Onwujekwe, R. Patel, E.A. Webb, D.A. Lawlor, and J.R. Hargreaves, Measuring socio-economic position for epidemiological studies in low-and middle-income countries: a methods of measurement in epidemiology paper. International Journal of Epidemiology, 2012. 41(3): p. 871–886.

33. McDade, T.W., J. Rutherford, L. Adair, and C.W. Kuzawa, Early origins of inflammation: microbial exposures in infancy predict lower levels of C-reactive protein in adulthood. Proceedings of the Royal Society B-Biological Sciences, 2010. 277(1684): p. 1129-1137.

34. McDade, T., J.N. Rutherford, L. Adair, and C. Kuzawa, Adiposity and pathogen exposure predict C-reactive protein in Filipino women. Journal of Nutrition, 2008. 138: p. 2442–7.

35. McDade, T., J.N. Rutherford, L. Adair, and C. Kuzawa, Population differences in C-reactive protein concentration and associations with adiposity: Comparing young adults in the Philippines and the U.S. American Journal of Clinical Nutrition, 2009. 89(4): p. 1237–1245.

36. McDade, T., M. Jones, G. Miller, J. Borja, M. Kobor, and C. Kuzawa, Birth weight and postnatal microbial exposures predict the distribution of peripheral blood leukocyte subsets in young adults in the Philippines. Journal of Developmental Origins of Health and Disease, 2018. 9(2): p. 198–207.

37. Brodin, P. and M.M. Davis, Human immune system variation. Nat Rev Immunol, 2017. 17(1): p. 21–29.

38. Yan, J., J.M. Greer, R. Hull, J.D. O’Sullivan, R.D. Henderson, S.J. Read, and P.A. McCombe, The effect of ageing on human lymphocyte subsets: comparison of males and females. Immunity and Ageing, 2010. 7: p. 1–10.

39. Hua, Y., J. Sun, X. Lou, W. Sun, and X. Kong, Monocyte-to-lymphocyte ratio predicts mortality and cardiovascular mortality in the general population. International Journal of Cardiology, 2023. 379: p. 118–126.

40. Phillips, A.C., D. Carroll, C.R. Gale, M. Drayson, and G.D. Batty, Lymphocyte cell counts in middle age are positively associated with subsequent all-cause and cardiovascular mortality. QJM: An International Journal of Medicine, 2010. 104(4): p. 319–324.

41. Franceschi, C. and J. Campisi, Chronic inflammation (inflammaging) and its potential contribution to age-associated diseases. Journals of Gerontology Series A: Biomedical Sciences and Medical Sciences, 2014. 69(Suppl_1): p. S4-S9.

42. McDade, T.W., Three common assumptions about inflammation, aging, and health that are probably wrong. Proceedings of the National Academy of Sciences, 2023. 120(51): p. e2317232120.

43. Gurven, M., H. Kaplan, J. Winking, D. Eid Rodriguez, S. Vasunilashorn, J.K. Kim, C. Finch, and E. Crimmins, Inflammation and infection do not promote arterial aging and cardiovascular disease risk factors among lean horticulturalists. PLoS One, 2009. 4(8): p. e6590.

