## Supplemental Table 1 for "Evaluation of a DNA methylation-based measure of chronic inflammation in two generations of adults in metropolitan Cebu, Philippines"

**Table S1.** Pearson correlations between study variables and DNAm-CRP for younger adults (N=1,665) and older females (N=1,070).

| | Young adults | Older females |
|---|---|---|
| age (years) | -0.0012 | -0.0541 |
| Daily smoker (0, 1) | **-0.095** | 0.0071 |
| Waist circumference (cm) | **0.1539** | **0.2769** |
| Household microbial environment (SD) | **0.0639** | -0.0231 |
| Household assets (items) | **-0.0559** | 0.0599 |
| Formal education (years) | **-0.0593** | 0.0219 |
| Infectious symptoms (0, 1) | **0.1169** | **0.0939** |
| CD4 T lymphocytes (%) | **-0.3153** | **-0.1415** |
| CD8 T lymphocytes (%) | **-0.3194** | **-0.2498** |
| B lymphocytes (%) | **-0.2361** | -0.0417 |
| Natural killer cells (%) | **-0.1984** | **-0.2003** |
| Monocytes (%) | 0.0252 | **0.0644** |
| Granulocytes (%) | **0.449** | **0.3038** |

Note: Bold text indicates correlations with $p<0.05$.
